# Interplay between mosaic chromosomal alterations and polygenic risk score increases risk of non-small cell lung cancer

**DOI:** 10.1101/2022.04.13.22273440

**Authors:** Na Qin, Congcong Chen, Liu Yang, Su Liu, Yuan Xie, Xianfeng Xu, Jun Zhou, Juncheng Dai, Guangfu Jin, Hongxia Ma, Cheng Wang, Hongbing Shen, Zhibin Hu

## Abstract

We investigated autosomal mosaic chromosomal alterations (mCAs) in 10,248 non-small cell lung cancer (NSCLC) cases and 9,298 cancer-free controls of Chinese ancestry. Mosaic loss and copy-neutral loss of heterozygosity were associated with an increased risk of NSCLC, while mosaic gain was associated with a decreased risk of NSCLC, especially those spanning telomeres. The increased cell fraction of mCAs was also correlated with an increasing NSCLC risk in the affected individuals. Both multiplicative and additive interactions were observed between polygenic risk score (PRS) and the presence of mosaic loss, where carriers of mosaic loss events with cell fractions ≥5% among the high genetic risk group had the greatest risk for developing NSCLC. These findings suggest that mCA events may act as a new endogenous indicator for risk of NSCLC and have the potential to be jointly used with PRS to optimize risk stratification of NSCLC.

## Introduction

Lung cancer is one of the most common incident cancers and the leading cause of cancer deaths worldwide^1^. The initiation and development of non-small cell lung cancer (NSCLC), the most common histology type of lung cancer, are believed to be influenced by a complex interplay between genetic architecture, aging, and environmental factors^2,3,4^. Genome-wide association studies (GWASs) have identified a number of inherited genetic loci for NSCLC^5,6^, and polygenic risk scores (PRS) constructed by combining these susceptibility loci have also been proved to be effective in quantifying individual risk of NSCLC^6,7^. However, the predictive performance of PRS differs among individuals depending on their age, lifestyles, or environmental exposures^8,9,10^, suggesting that the effect of heritable factors can be modified by non-heritable environmental risk factors.

In addition to tobacco smoking, a well-known risk factor for lung cancer^11^, recent evidence has linked several other environmental exposures to NSCLC risk, such as ambient air pollutants^9,12^, inhalable agents^13^, hazardous chemicals^14,15,16,17^ and unfavorable lifestyles^11,18,19,20^; however, the evaluation of associations between environmental factors and cancer risk is still challenging, as environmental exposures are temporally dynamic and difficult to measure accurately^21,22,23^, letting alone individuals who may have varied vulnerability and susceptibility in response to the same exposure^24,25^. Thus, an endogenous biomarker that could reflect internal effect of external environmental agents on biological capability is warranted to help yield efficient prediction of cancer risk.

Mosaic chromosomal alterations (mCAs) are large clonal structural somatic alterations detected in the whole blood-derived DNA, which are characterized by whole-chromosomal or sub-chromosomal level changes, including gain, loss, and copy-neutral loss of heterozygosity (CN-LOH). In addition to the crucial role of aging in the exponential increase of mCAs^26,27^, several other factors have also been reported to help shape the mCA landscapes, such as ambient air pollutants^28^, hazardous chemicals^29,30^, radiation^31^, and unhealthy lifestyles^32,33,34,35^, suggesting that mCAs can be viewed as an internal genomic signature associated with both internal and external exposures^36^. In the past decades, large population-based studies have established the effect of chromosomal mosaicism on early human embryogenesis^37,38^, birth defects^39,40^ and a broad range of adult diseases^41,42^ and cancers^26,27,31,43,44,45^. However, little is known about the type, frequency, and effect of acquired chromosomal anomalies to be associated with the development of NSCLC as well as their interplay with genetic factors.

Thus, in the current study, by using genotype array data of 10,248 NSCLC patients and 9,298 cancer-free controls of Chinese anscetry, we intend to systematically describe the landscape of mCAs in Chinese populations, investigate associations of detectable mCAs with risk of NSCLC, and examine their interplay with genetic factors (i.e., PRS).

## Methods

### Study populations

A total of 19,546 participants of Han Chinese ancestry were included in the current study, including 10,248 NSCLC cases and 9,298 cancer-free controls. The study population was derived from our recently published GWAS^6^ which included 4,149 cases and 3,198 controls from Nanjing and Shanghai, 2,155 cases and 2,035 controls from Beijing and Tianjin, and 3,944 cases and 4,065 controls from Guangdong. The demographics of these subjects were described in **Supplementary Table 1**. Of the included NSCLC cases, 6,839 were lung adenocarcinoma and 2,704 were lung squamous cell carcinomas. All lung cancer cases were histologically confirmed as new NSCLC cases by at least two local pathologists and were free of chemotherapy or radiotherapy before diagnosis. The included cancer-free controls were selected from a local community-based screening program for non-infectious diseases in Chinese populations. Written informed consent was obtained from all of the included subjects, and the study was approved by the Institutional Review Board (IRB) at Nanjing Medical University.

### Genotyping and quality control

Blood-derived DNA of the included participants was genotyped with the Infinium Global Screening Array (GSA, version 1.0) that included 700,078 markers. Duplicate markers or SNPs were excluded from further analysis, if (1) with call rate <95%, (2) not in Hardy-Weinberg equilibrium (HWE, *P*<10^−7^ in controls or *P*<10^−12^ in NSCLC cases), (3) differed in frequency from the 1000 Genomes Project dataset, or (4) falling within segmental duplications with low divergence (<2%). Variant-level quality control was performed for each subpopulation independently. Finally, 581,940 (in Nanjing GSA GWAS), 581,252 (in Beijing GSA GWAS), and 605,292 (in Zhongshan GSA GWAS) variants on autosome chromosomes were included in the following analyses.

### Haplotype phasing and detection of mCAs

The detection of mCAs was conducted on raw IDAT files from the Illumina GSA. Firstly, the genotype clustering was performed using the Illumina GenCall algorithm, and the resulting GTC files were converted to VCF files with the BCFtools gtc2vcf plugin (https://github.com/freeseek/gtc2vcf). The log2 R ratio (LRR) and B-allele frequency (BAF) values were used to estimate the total and relative allelic intensities, respectively. Then, SHAPEIT4 (version 4.2.1)^46^ was used for phasing across the three sample sets with default parameters, and the phased genotypes were ligated across overlapping windows using BCFtools concat plugin. Finally, the mCA calling was performed with Mosaic Chromosomal Alterations (MoChA)^43,47^ by leveraging long-range phase information to detect allelic imbalances across contiguous genomic segments. The identified mCAs were classified into copy number gain, loss, and CN-LOH based on the LRR and BAF values, and those that could not be assigned to any of the three types were designated as ‘undetermined’ (**Supplementary Fig. 1**).

Following previous studies, to filter out possible constitutional or inherited duplications, we excluded mCA events of length >0.5 Mb with relative coverage ≥2.5 or Bdev ≥0.1, and mCAs of length >5 Mb with Bdev ≥0.15. mCA events within the major histocompatibility complex (MHC) region (chr6:28,477,797–33,448,354, hg19) were further excluded due to the known propensity to call false-positive mosaic CN-LOH events within this locus.

The chromosomal location of mCA events was defined as follows: (1) events that only occurred around telomeric ends (±1 Mb from chromosome ends) were defined as telomere events, (2) events that crossed the centromere were defined as centromeric, (3) events that spanned an entire chromosome were defined as whole chromosome events, and (4) all other events were defined as interstitial.

### Construction of polygenetic risk score

Based on all previously reported susceptibility SNPs (i.e., 81 SNPs in 40 loci) for lung cancer at a genome-wide significance level, our previous study constructed a PRS with 19 risk variants that are specific for Chinese populations^6^. Detailed information for the 19 SNPs included in the PRS was extracted from our previous study as described in **Supplementary Table 2**.

In the current study, the PRS was generated by summing all the 19 variants with the following formula: 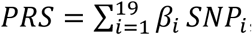, where *β*_i_ is the estimated weight (lnOR) of the *i*th SNP derived from our previous study^6^ and *SNP*_i_ is the genotype dosage of each risk allele for each variant. The PRS was categorized as low (the first quartile), intermediate (25%∼75%) and high (the upper quartile) genetic risk (percentages were based on the distribution of the PRS among the cancer-free controls).

### Statistical analysis

Logistic regression analysis was applied to evaluate the association between the presence of mCAs or specific mCA types and risk of NSCLC with adjustment for age, sex, smoking pack-years, DNA source, and ten principal components, in which odds ratios (ORs) and standard errors (SEs) were calculated. The multiplicative interaction was quantified by including a product term of genetic risk and the presence of mCA events in the model. The additive interaction was measured by calculating the relative excess risk due to interaction (RERI) and the attributable proportion because of the interaction (AP) based on coefficients of the product term. A Wilcoxon rank-sum test was used to assess the difference in the magnitude of different mCA types. All the reported *P* values were two-sided. All the analyses were performed using the R software (version 3.5.1, R Project for Statistical Computing).

## Results

### Characterization of the detected mCA events

Among the 10,248 NSCLC cases and 9,298 controls, we detected a total of 956 autosomal mCAs in 747 individuals (a prevalence of 3.82%), including 572 mCAs in 418 NSCLC cases (a prevalence of 4.08%) and 384 mCAs in 329 cancer-free controls (a prevalence of 3.54%) (**Table 1**). Of the mCA carriers, 657 had only one, 46 two and 44 at least three mCA events (**Fig. 1a**). Of the detected mCA events, the most frequent type was copy loss (36.40%, 348/956), whereas copy gain and CN-LOH constituted 23.01% (220/956) and 28.87% (276/956) of the mCA events, respectively (**Table 1**). In addition, most (54.71%, 523/956) of the mosaic chromosomal events began at a telomere and extended across some portion of the chromosomal arm, while only a small proportion (4.08%, 39/956) of mCA events spanned the entire chromosome, and 1.05% (10/956) crossed the centromere. The remaining mCA events were interstitial (40.17%, 384/956), spanning neither telomere nor centromere, and the chromosomal distribution of mCAs varied across different types (**Supplementary Fig. 2**).

**Fig. 1.**
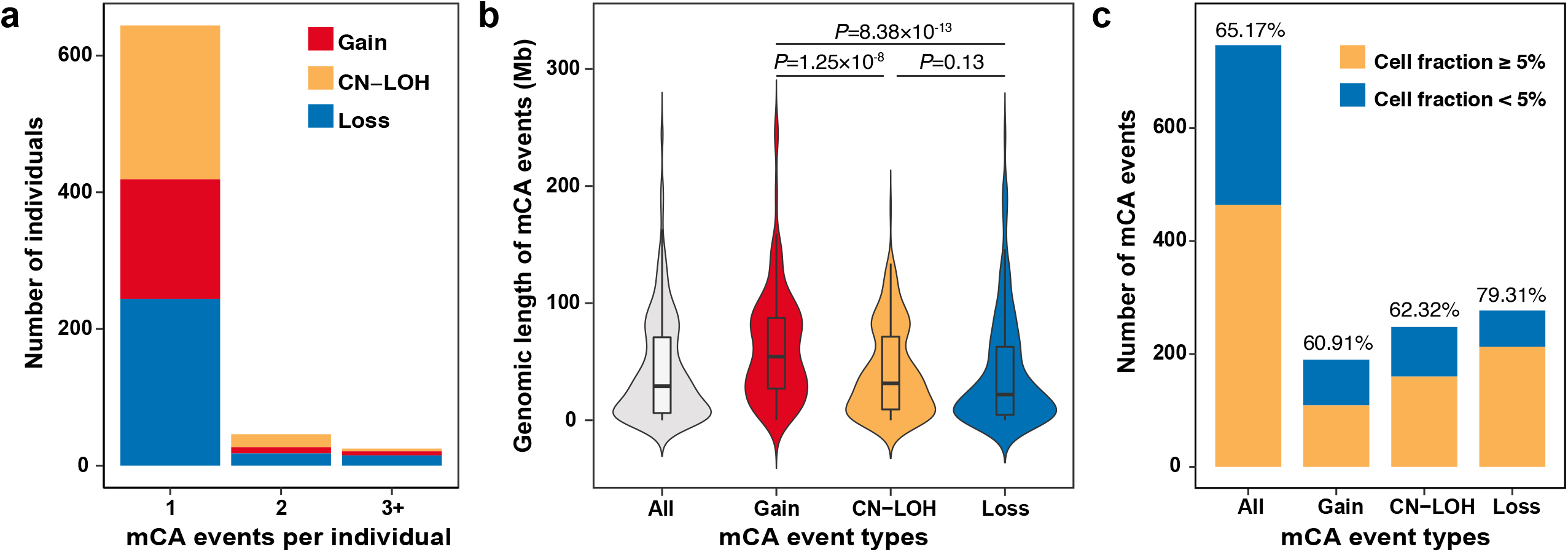
Distributional properties of the detected autosomal mosaic chromosomal alterations (mCAs). (a) Distribution of the number of mCAs in each individual. Individuals with mosaic chromosomal gain, loss, or copy-neutral loss of heterozygosity (CN-LOH) are illustrated by different colors. (b) The genomic magnitude of mCAs by copy number alteration types. (c) Distribution of the number and proportion of mCAs with cell fraction ≥5%.

**Table 1.** Counts and prevalence of mosaic chromosomal alterations (mCAs) by event type and genomic location.

|  | Overall populations (N=19,546) |  | NSCLC cases (N=10,248) |  | Cancer-free controls (N=9,298) |  |
| --- | --- | --- | --- | --- | --- | --- |
|  | No. of events<br>(no. of samples,<br>prevalence) | Proportion | No. of events<br>(no. of samples,<br>prevalence) | Proportion | No. of events<br>(no. of samples,<br>prevalence) | Proportion |
| **Total** | 956 (747, 3.82%) | 100.00% | 572 (418, 4.08%) | 100.00% | 384 (329, 3.54%) | 100.00% |
| **Copy number change** |  |  |  |  |  |  |
| Gain | 220 (190, 0.97%) | 23.01% | 90 (79, 0.77%) | 15.73% | 130 (111, 1.19%) | 33.85% |
| Loss | 348 (277, 1.42%) | 36.40% | 248 (179, 1.75%) | 43.36% | 100 (98, 1.05%) | 26.04% |
| CN-LOH | 276 (248, 1.27%) | 28.87% | 170 (153, 1.49%) | 29.72% | 106 (95, 1.02%) | 27.60% |
| Undetermined | 112 (101, 0.52%) | 11.72% | 64 (55, 0.54%) | 11.19% | 48 (46, 0.49%) | 12.50% |
| **Genomic location** |  |  |  |  |  |  |
| Whole chromosome | 39 (31, 0.16%) | 4.08% | 28 (21, 0.20%) | 4.90% | 11 (10, 0.11%) | 2.86% |
| Telomeric p | 164 (158, 0.81%) | 17.15% | 90 (87, 0.85%) | 15.73% | 74 (71, 0.76%) | 19.27% |
| Telomeric q | 359 (338, 1.73%) | 37.55% | 202 (187, 1.82%) | 35.31% | 157 (151, 1.62%) | 40.89% |
| Interstitial | 384 (302, 1.55%) | 40.17% | 245 (184, 1.80%) | 42.83% | 139 (118, 1.27%) | 36.20% |
| Centromeric | 10 (10, 0.05%) | 1.05% | 7 (7, 0.07%) | 1.22% | 3 (3, 0.03%) | 0.78% |

Abbreviations: CN-LOH, copy-neutral loss of heterozygosity; NSCLC, non-small cell lung cancer.

The size of detected mCA events was broadly consistent with those found in previous studies^41,45^, ranging from 16.97 kb to 249.25 Mb (median, 29.14 Mb) (**Fig. 1b**). The average magnitude of mosaic copy number gain events was significantly longer than that of mosaic CN-LOH (54.33 vs 31.57 Mb, *P*=1.25×10^−8^) and loss events (54.33 vs 21.99 Mb, *P*=8.38×10^−13^) (**Fig. 1b**). The estimated proportion of cells harboring an mCA event was widely distributed, ranging from 1.03% to 67.73% with a median of 7.53%, where a substantial number (N=623, 65.17%) of mCAs was seen with a cell fraction of ≥5%, particularly for mosaic loss events (N=276, 79.31%) (**Fig. 1c**). More than half of the mosaic gain (N=134, 60.91%) and CN-LOH (N=172, 62.32%) abnormalities also had a relatively larger cell fraction (**Fig. 1c**).

### Demographic associations with mCAs

As age is a known risk factor for mCAs, we also observed an increased prevalence of autosomal mCAs with age in both NSCLC cases and cancer-free controls (**Fig. 2, Supplementary Table 3**). Interestingly, the proportion of mCA carriers was even higher among NSCLC cases than controls from age under 50 (2.61% *vs* 1.78%) to age greater than 80 (13.58% *vs* 7.97%) (**Fig. 2**), especially for the mosaic loss events (**Fig. 2, Supplementary Fig. 3**). No statistical association was observed for age at DNA collection with the magnitude or the clone size of mCA events (*P*>0.05) (**Supplementary Fig. 4**).

**Fig. 2.**
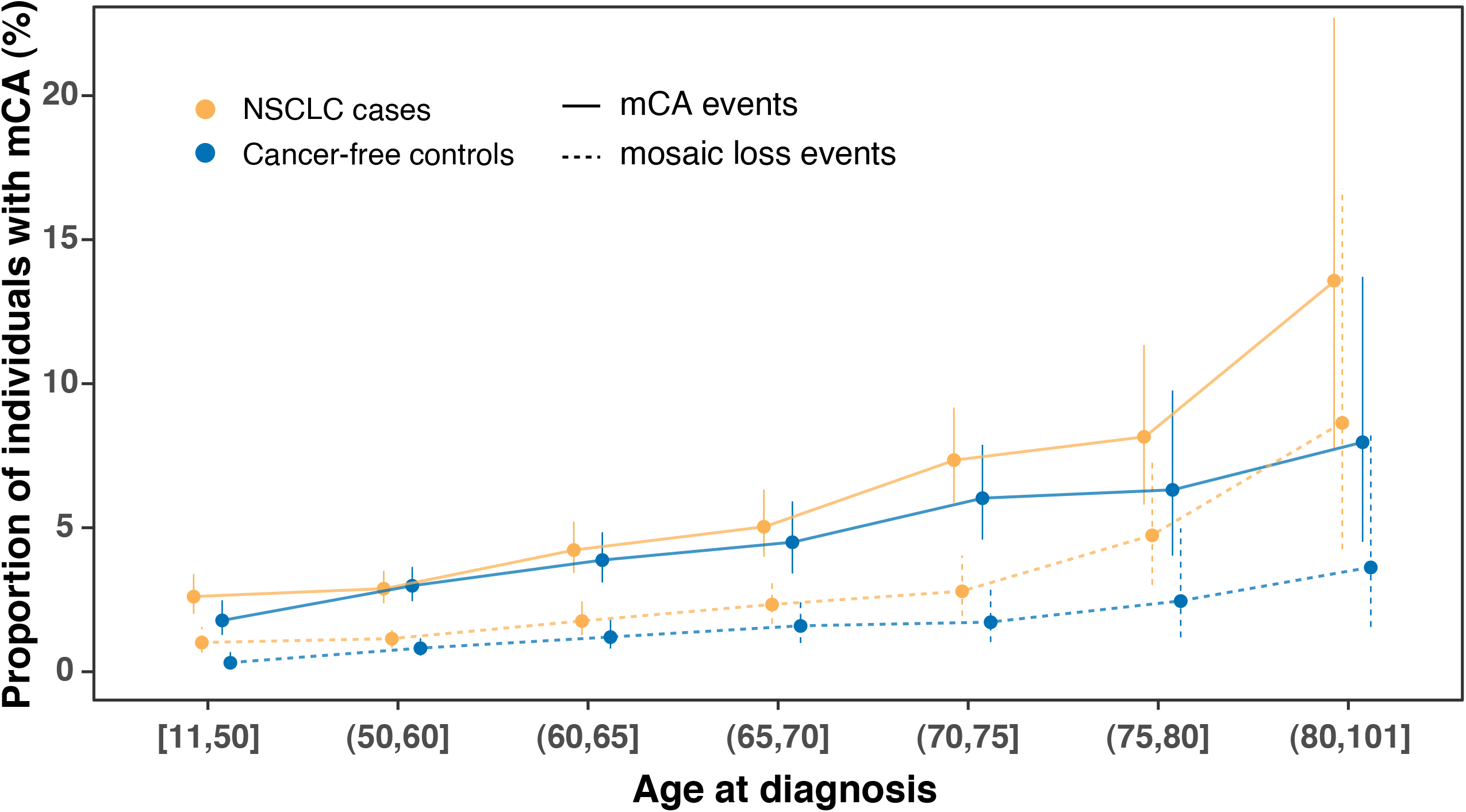
Proportion of individuals with the detected mosaic chromosomal alterations (mCAs) increases with age.

To evaluate the association of other common demographic factors with the incidence of mCAs, we compared the prevalence of mCAs among subgroups by sex and smoking status (**Supplementary Table 3**). Most mosaic gain events were enriched in males (1.23% *vs* 0.53%, *P*=4.73×10^−7^) and smokers (1.20% *vs* 0.78%, *P*=3.34×10^−3^), and mosaic CN-LOH events also tended to affect smokers (1.42% *vs* 1.14%, *P*=0.083), although not reaching a significance level. When age, sex, and smoking status were included in the same multivariable model, age still showed a significant association with the mCA frequency (OR=1.99, 95% CI=1.71-2.33; *P*=3.35×10^−18^), while males carried a significantly higher burden of mosaic gain events than females (OR=2.27, 95% CI=1.49-3.45; *P*=1.25×10^−4^) and smokers had significantly more mosaic CN-LOH events than life-long non-smokers (OR=1.46, 95% CI=1.04-2.05; *P*=0.031) (**Supplementary Table 4**).

### Associations of autosomal mCAs with lung cancer risk

As the prevalence of mCA carriers among NSCLC cases was higher than that of controls across different age groups, we then evaluated the relationship between detectable autosomal mCAs and risk of NSCLC. We identified that the presence of mCA events was associated with a significantly elevated risk of NSCLC (OR=1.17, 95% CI=1.00-1.37; *P*=0.045) after adjustment for age, sex, smoking pack-years, DNA source, and ten principal components (**Fig. 3a**). Subgroup analysis by event type revealed that mosaic copy number loss had the strongest effect on NSCLC risk (OR=1.77, 95% CI=1.37-2.30; *P*=1.43×10^−5^), and mosaic CN-LOH also contributed to an elevated risk of NSCLC (OR=1.34, 95% CI=1.02-1.77; *P*=0.033) (**Fig. 3a**). In contrast, mosaic copy number gain was found to have a protective effect on NSCLC (OR=0.69, 95% CI=0.51-0.94; *P*=0.019) (**Fig. 3a**), and the effect was primarily observed in females (OR=0.34, 95% CI=0.16-0.72; *P*=4.87×10^−3^ for females and OR=0.84, 95% CI=0.60-1.19; *P*=0.333; *P*_Het_=0.032 for males) (**Supplementary Table 5**).

**Fig. 3.**
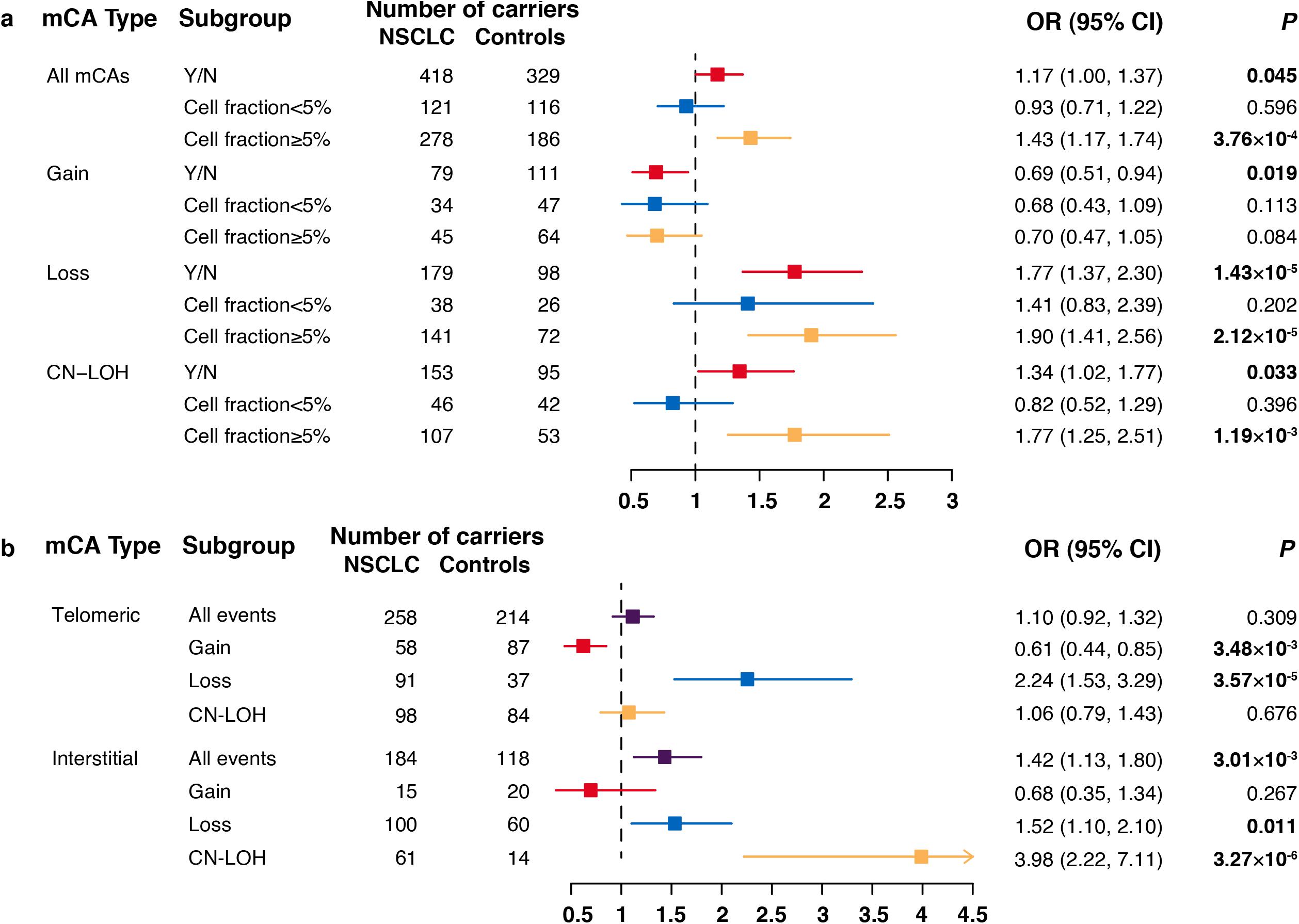
Associations of mosaic chromosomal alterations (mCAs) with risk of non-small cell lung cancer (NSCLC). (a) Odds ratios (ORs) for the association between risk of NSCLC and mCAs by copy number alteration types. mCAs with cell fractions less than 5% or not are illustrated by different colors. Error bars indicate 95% confidence intervals (CIs). A total of 55 mCA events designated as ‘Undetermined’ failed in cell fraction estimation. (b) ORs for the association between risk of NSCLC and mCAs by genomic locations. Mosaic chromosomal gain, loss, or copy-neutral loss of heterozygosity (CN-LOH) are illustrated by different colors. Error bars indicate 95% CIs.

Then, we divided the mCAs according to their genomic locations (**Fig. 3b, Supplementary Table 6**) and observed that most (71.88%) of interstitial events were mosaic loss and CN-LOH, which all contributed to an elevated risk of NSCLC (mosaic loss: OR=1.52, 95% CI=1.10-2.10; *P*=0.011; mosaic CN-LOH: OR=3.98, 95% CI=2.22-7.11; *P*=3.27×10^−6^). Most of the mosaic gain events (77.00%) span across telomere regions, and had a protective effect on NSCLC (OR=0.61, 95% CI=0.44-0.85; *P*=3.48×10^−3^). As the effect of mosaic abnormalities on NSCLC risk may vary by cell fraction, we divided the mCAs according to their cell fractions, and identified that the presence of mCAs with large cell fractions (≥5%) had a much stronger association with the risk of NSCLC (OR=1.43 *vs* 0.93; *P*_Het_=0.012; **Fig. 3a**). In addition, we identified that the risk of NSCLC was much more strongly dependent on cell fractions (**Supplementary Table 7**).

### Interaction between polygenic risk score and mCAs

Among the major interests in the current study is the joint effect of the presence of mCAs and genetic factors. To determine this, we first constructed the PRS of NSCLC with the 19 SNPs reported in our previous study (**Supplementary Table 2**). NSCLC cases tended to have a higher PRS (genetic risk) than those cancer-free controls (**Supplementary Fig. 5**). Interestingly, there was a synergetic effect of genetic factor (i.e., PRS) and mosaic loss events on risk of NSCLC in a dose-response manner (*P* for multiplicative interaction analysis =0.030) (**Supplementary Table 8**), where the overall risk of NSCLC increased as both PRS and the occurrence of mosaic loss events increased (**Fig. 4**). Specifically, compared to non-carriers of mosaic loss event with low genetic risk, carriers of mosaic loss abnormalities with a high genetic risk had the highest risk of developing NSCLC (OR=6.16, 95% CI=3.66-10.36; *P*=7.35×10^−12^) (**Fig. 4**), and the effect greatly increased in those who carry mosaic loss with cell fractions ≥5% (OR=7.64, 95% CI=4.01-14.56; *P*=6.25×10^−10^) (**Fig. 4**).

**Fig. 4.**
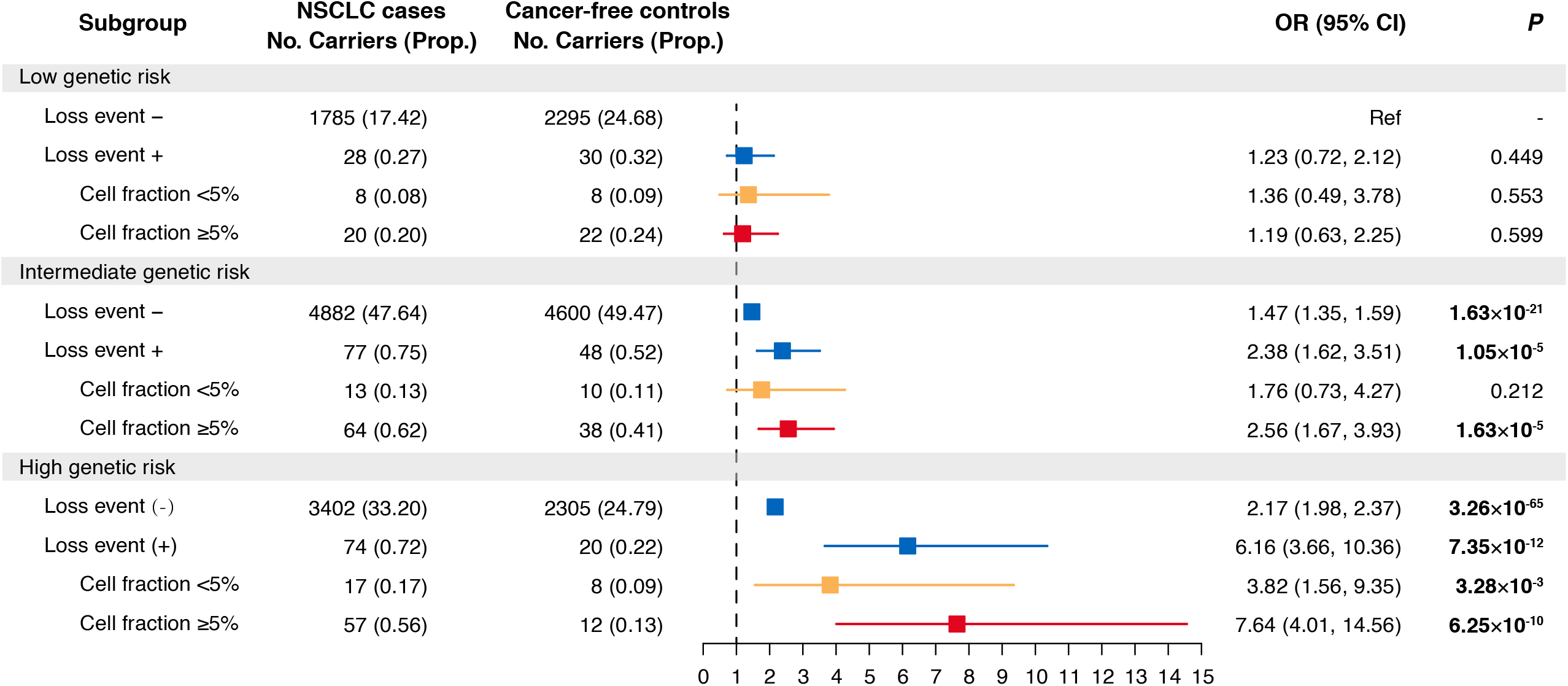
Risk of non-small cell lung cancer (NSCLC) according to the detected mosaic chromosomal loss and genetic categories. The associations between mosaic loss abnormalities and risk of NSCLC according to genetic categories (i.e., polygenic risk score). Abbreviations: OR, odds ratio; CI, confidence interval.

Significant additive interaction was also found between PRS and the presence of expanded mosaic loss (**Supplementary Table 8**): for carriers of mosaic loss events with cell fractions ≥5% among the highest genetic risk group, the RERI was 5.26 (95% CI=1.54-12.10), which suggests that there would be a 5.26-fold relative excess risk because of the additive interaction, accounting for 69% (31%-80%) of the risk in those participants who had both mosaic loss events and high genetic risk.

## Discussion

In the current study, we comprehensively delineated clonal hematopoiesis in terms of copy number alterations among 19,546 individuals of Chinese ancestry and provided novel evidence for associations between mCAs and risk of NSCLC. We found that mosaic chromosomal abnormalities, especially mosaic loss of copy number, contributed to an elevated risk of NSCLC and that the effect increased as the cell fraction of the events increased. More importantly, we demonstrated a joint effect of mosaic copy number alterations and genetic risk, where the greatest relative increase of NSCLC risk was observed among those with expanded mosaic loss alterations and the highest level of genetic risk. Our findings provide novel evidence that mCA may serve as an endogenous genomic indicator for risk of lung cancer and have the potential to be jointly used with PRS to optimize the risk stratification for lung cancer.

Mosaic chromosomal alterations detected from blood-derived DNA are a group of somatic alterations that emerge in the period of early embryonic development and at the following stages of the lifespan^48,49^. Because this type of structural genomic alterations simultaneously spans dozens to hundreds or even thousands of genes, they have long been recognized for their necessity in hematological malignancy and mortality^43,31,44^. Recently, several studies proposed that mCAs could also be a possible source of unexplained susceptibility of solid cancers^26,27,50^; however, these studies have limited sample sizes not enough for cancer-specific analyses nor for copy number alteration type-specific (i.e., gain, loss, and CN-LOH) analyses. Our study demonstrated different effects of mCA types on NSCLC risk: while mosaic copy number loss and CN-LOH conferred an increased risk of NSCLC, the mosaic gain of copy numbers appeared to have a protective effect on NSCLC. Large copy number loss distributed across the genome was widely accepted as a risk factor of multiple cancers, no matter in the germline or somatic cells^51,52,53^. The mosaic gain, which primarily encompasses the telomere region, may help preserve genome stability and prevent the development of cancer.

Polygenic risk scores (PRSs) have advanced substantially in recent years and are widely used as an informative genetic measure for discriminating subpopulations at high risk of site-specific cancers^6,7,54,55,56,57,58,59,60,61,62,63^. Our recent work also constructed a PRS to stratify risk of lung cancer^6^. However, because many cancer types involve both environmental factors and genetic susceptibility, PRS in combination with other known risk factors will further improve risk prediction in the clinics. Here, we provided the first evidence, to the best of our knowledge, that mosaic loss of copy number synergistically with genetic factors increased risk of NSCLC and that approximately 69% of such an increased NSCLC risk could be attributed to their additive interaction, suggesting that incorporating mCAs into the existing PRS prediction model may contribute to a much improved predictive efficiency. In clinical practice, mCA may be expected as an ideal biomarker for risk assessment. Firstly, it could be a byproduct of the SNP array used for the PRS construction with no more tests^43^. Secondly, mCA can be monitored throughout life and provide a dynamic risk assessment coupled with PRS, given that mCAs provide a lifetime tracking of endogenous genomic alterations in response to environmental exposures^26,64^. With the development of electronic health records and the increasing availability of population biobanks, these findings should facilitate these resources to be effectively used to improve the precision risk prediction of lung cancer.

Although our results provide novel insights into associations of genomic mCAs with lung cancer, the interpretation of our findings needs to be considered within the following limitations. First, although this is the largest study for evaluating lung cancer risk associated with mCAs among Chinese populations, the number of detected mCA events is still not large enough to further investigate effect modification of genetic or environmental factors by such genomic alterations. In addition, the modest number of mCAs we obtained also precluded us from investigating properties of mCAs, such as different magnitude scales and chromosomal distribution with the incidence of lung cancer. Finally, although we observed a synergistic effect of genetic factors and mosaic copy number loss on risk of NSCLC, the case-control design used in the current study could not provide precise estimates of the risk. Further large-scale cohort studies are warranted to replicate our findings and refine the risk estimates.

In conclusion, the current study demonstrates that mosaic chromosomal alterations contribute to an increased risk of NSCLC, which greatly substantiates the effect of genetic factors on lung cancer and also provides novel insights into the precise prediction of lung cancer.

## Data Availability

All the detected mosaic chromosomal alteration events and genotypic information for the 19 SNPs used for the calculation of polygenic risk score will be included as Supplemental Data before publication.

## Author Contributions

ZH, CW, and NQ initiated and conceived the study. ZH, HS, GJ, HM, and JD assisted in data collection and recruitment of study participants. CC and LY assisted in sample collection and processing with SL, YX, YX, and JZ. NQ and CW developed the analysis plan and performed the bioinformatics/statistical analyses. NQ and CW wrote the first draft of the manuscript. All authors contributed to data interpretation and writing and approved the final manuscript for publication.

## Acknowledgments

The authors would like to thank all the research staffs and students who participated in this work. This work was supported by the National Natural Science of China (81820108028, 81521004, 81922061, 82103926, 82072579), Research Unit of Prospective Cohort of Cardiovascular Diseases and Cancer, Chinese Academy of Medical Sciences (2019RU038), Natural Science Foundation of Jiangsu Province (BK20210534), and Postdoctoral Science Foundation of China (2021M691636).

## References

1. Sung H, et al. Global Cancer Statistics 2020: GLOBOCAN Estimates of Incidence and Mortality Worldwide for 36 Cancers in 185 Countries. CA: a cancer journal for clinicians 71, 209–249 (2021).

2. Molina JR, Yang P, Cassivi SD, Schild SE, Adjei AA. Non-small cell lung cancer: epidemiology, risk factors, treatment, and survivorship. Mayo Clinic proceedings 83, 584–594 (2008).

3. Meiners S, Eickelberg O, Konigshoff M. Hallmarks of the ageing lung. Eur Respir J 45, 807–827 (2015).

4. White MC, Holman DM, Boehm JE, Peipins LA, Grossman M, Henley SJ. Age and cancer risk: a potentially modifiable relationship. Am J Prev Med 46, S7–15 (2014).

5. Bosse Y, Amos CI. A Decade of GWAS Results in Lung Cancer. Cancer epidemiology, biomarkers & prevention : a publication of the American Association for Cancer Research, cosponsored by the American Society of Preventive Oncology 27, 363–379 (2018).

6. Dai J, et al. Identification of risk loci and a polygenic risk score for lung cancer: a large-scale prospective cohort study in Chinese populations. Lancet Respir Med 7, 881–891 (2019).

7. Hung RJ, et al. Assessing Lung Cancer Absolute Risk Trajectory Based on a Polygenic Risk Model. Cancer Res 81, 1607–1615 (2021).

8. Jiang X, Holmes C, McVean G. The impact of age on genetic risk for common diseases. PLoS Genet 17, e1009723 (2021).

9. Huang Y, et al. Air Pollution, Genetic Factors, and the Risk of Lung Cancer: A Prospective Study in the UK Biobank. Am J Respir Crit Care Med 204, 817–825 (2021).

10. Zhu M, et al. Genetic Risk for Overall Cancer and the Benefit of Adherence to a Healthy Lifestyle. Cancer Res 81, 4618–4627 (2021).

11. Islami F, et al. Proportion and number of cancer cases and deaths attributable to potentially modifiable risk factors in the United States. CA Cancer J Clin 68, 31–54 (2018).

12. Turner MC, et al. Outdoor air pollution and cancer: An overview of the current evidence and public health recommendations. CA Cancer J Clin, (2020).

13. Mu L, et al. Indoor air pollution and risk of lung cancer among Chinese female non-smokers. Cancer Causes Control 24, 439–450 (2013).

14. Roscoe RJ, Steenland K, Halperin WE, Beaumont JJ, Waxweiler RJ. Lung cancer mortality among nonsmoking uranium miners exposed to radon daughters. JAMA 262, 629–633 (1989).

15. Chen CL, et al. Ingested arsenic, cigarette smoking, and lung cancer risk: a follow-up study in arseniasis-endemic areas in Taiwan. JAMA 292, 2984–2990 (2004).

16. Brims FJH, et al. Pleural Plaques and the Risk of Lung Cancer in Asbestos-exposed Subjects. Am J Respir Crit Care Med 201, 57–62 (2020).

17. Li N, et al. Association of 13 Occupational Carcinogens in Patients With Cancer, Individually and Collectively, 1990-2017. JAMA Netw Open 4, e2037530 (2021).

18. Chen W, et al. Disparities by province, age, and sex in site-specific cancer burden attributable to 23 potentially modifiable risk factors in China: a comparative risk assessment. Lancet Glob Health 7, e257–e269 (2019).

19. Alpha-Tocopherol BCCPSG. The effect of vitamin E and beta carotene on the incidence of lung cancer and other cancers in male smokers. N Engl J Med 330, 1029–1035 (1994).

20. Omenn GS, et al. Effects of a combination of beta carotene and vitamin A on lung cancer and cardiovascular disease. N Engl J Med 334, 1150–1155 (1996).

21. Feng J, Liu T, Qin B, Zhang Y, Liu XS. Identifying ChIP-seq enrichment using MACS. Nature protocols 7, 1728–1740 (2012).

22. Beyond Space (As We Knew It): Toward Temporally Integrated Geographies of Segregation, Health, and Accessibility. Annals of the Association of American Geographers 103, (2013).

23. Park YM, Kwan M-P. Individual exposure estimates may be erroneous when spatiotemporal variability of air pollution and human mobility are ignored. Health and Place 43, (2017).

24. Grandjean P. Individual susceptibility in occupational and environmental toxicology. Toxicol Lett 77, 105–108 (1995).

25. Raza A, Dahlquist M, Lind T, Ljungman PLS. Susceptibility to short-term ozone exposure and cardiovascular and respiratory mortality by previous hospitalizations. Environ Health 17, 37 (2018).

26. Laurie CC, et al. Detectable clonal mosaicism from birth to old age and its relationship to cancer. Nat Genet 44, 642–650 (2012).

27. Jacobs KB, et al. Detectable clonal mosaicism and its relationship to aging and cancer. Nat Genet 44, 651–658 (2012).

28. Wong JYY, et al. Outdoor air pollution and mosaic loss of chromosome Y in older men from the Cardiovascular Health Study. Environ Int 116, 239–247 (2018).

29. Bai Y, et al. Effects of polycyclic aromatic hydrocarbons and multiple metals co-exposure on the mosaic loss of chromosome Y in peripheral blood. J Hazard Mater 414, 125519 (2021).

30. Liu Y, et al. Polycyclic aromatic hydrocarbons exposure and their joint effects with age, smoking, and TCL1A variants on mosaic loss of chromosome Y among coke-oven workers. Environ Pollut 258, 113655 (2020).

31. Gao T, et al. Interplay between chromosomal alterations and gene mutations shapes the evolutionary trajectory of clonal hematopoiesis. Nat Commun 12, 338 (2021).

32. Dumanski JP, et al. Mutagenesis. Smoking is associated with mosaic loss of chromosome Y. Science 347, 81–83 (2015).

33. Dawoud AAZ, Tapper WJ, Cross NCP. Clonal myelopoiesis in the UK Biobank cohort: ASXL1 mutations are strongly associated with smoking. Leukemia 34, 2660–2672 (2020).

34. Zhou W, et al. Mosaic loss of chromosome Y is associated with common variation near TCL1A. Nat Genet 48, 563–568 (2016).

35. Qin N, et al. Association of Mosaic Loss of Chromosome Y with Lung Cancer Risk and Prognosis in a Chinese Population. J Thorac Oncol 14, 37–44 (2019).

36. Dai X, Guo X. Decoding and rejuvenating human ageing genomes: Lessons from mosaic chromosomal alterations. Ageing Res Rev 68, 101342 (2021).

37. Vanneste E, et al. Chromosome instability is common in human cleavage-stage embryos. Nat Med 15, 577–583 (2009).

38. Yang X, et al. Developmental and temporal characteristics of clonal sperm mosaicism. Cell 184, 4772–4783 e4715 (2021).

39. Conlin LK, et al. Mechanisms of mosaicism, chimerism and uniparental disomy identified by single nucleotide polymorphism array analysis. Hum Mol Genet 19, 1263–1275 (2010).

40. Ballif BC, et al. Detection of low-level mosaicism by array CGH in routine diagnostic specimens. Am J Med Genet A 140, 2757–2767 (2006).

41. Sherman MA, et al. Large mosaic copy number variations confer autism risk. Nat Neurosci 24, 197–203 (2021).

42. Zekavat SM, et al. Hematopoietic mosaic chromosomal alterations increase the risk for diverse types of infection. Nat Med 27, 1012–1024 (2021).

43. Loh PR, et al. Insights into clonal haematopoiesis from 8,342 mosaic chromosomal alterations. Nature 559, 350–355 (2018).

44. Terao C, et al. Chromosomal alterations among age-related haematopoietic clones in Japan. Nature 584, 130–135 (2020).

45. Saiki R, et al. Combined landscape of single-nucleotide variants and copy number alterations in clonal hematopoiesis. Nat Med 27, 1239–1249 (2021).

46. Delaneau O, Zagury JF, Robinson MR, Marchini JL, Dermitzakis ET. Accurate, scalable and integrative haplotype estimation. Nature communications 10, 5436 (2019).

47. Loh PR, Genovese G, McCarroll SA. Monogenic and polygenic inheritance become instruments for clonal selection. Nature 584, 136–141 (2020).

48. Youssoufian H, Pyeritz RE. Mechanisms and consequences of somatic mosaicism in humans. Nat Rev Genet 3, 748–758 (2002).

49. Thorpe J, Osei-Owusu IA, Avigdor BE, Tupler R, Pevsner J. Mosaicism in Human Health and Disease. Annu Rev Genet 54, 487–510 (2020).

50. Loftfield E, Zhou W, Yeager M, Chanock SJ, Freedman ND, Machiela MJ. Mosaic Y Loss Is Moderately Associated with Solid Tumor Risk. Cancer Res 79, 461–466 (2019).

51. Zack TI, et al. Pan-cancer patterns of somatic copy number alteration. Nat Genet 45, 1134–1140 (2013).

52. Li Y, et al. Patterns of somatic structural variation in human cancer genomes. Nature 578, 112–121 (2020).

53. Huang KL, et al. Pathogenic Germline Variants in 10,389 Adult Cancers. Cell 173, 355–370 e314 (2018).

54. Mavaddat N, et al. Polygenic Risk Scores for Prediction of Breast Cancer and Breast Cancer Subtypes. Am J Hum Genet 104, 21–34 (2019).

55. Mars N, et al. The role of polygenic risk and susceptibility genes in breast cancer over the course of life. Nat Commun 11, 6383 (2020).

56. Liu C, et al. Generalizability of Polygenic Risk Scores for Breast Cancer Among Women With European, African, and Latinx Ancestry. JAMA Netw Open 4, e2119084 (2021).

57. Kachuri L, et al. Pan-cancer analysis demonstrates that integrating polygenic risk scores with modifiable risk factors improves risk prediction. Nat Commun 11, 6084 (2020).

58. Liyanarachchi S, et al. Assessing thyroid cancer risk using polygenic risk scores. Proc Natl Acad Sci U S A 117, 5997–6002 (2020).

59. Frampton MJ, et al. Implications of polygenic risk for personalised colorectal cancer screening. Ann Oncol 27, 429–434 (2016).

60. Thomas M, et al. Genome-wide Modeling of Polygenic Risk Score in Colorectal Cancer Risk. Am J Hum Genet 107, 432–444 (2020).

61. Plym A, et al. Evaluation of a Multiethnic Polygenic Risk Score Model for Prostate Cancer. J Natl Cancer Inst, (2021).

62. Jin G, et al. Genetic risk, incident gastric cancer, and healthy lifestyle: a meta-analysis of genome-wide association studies and prospective cohort study. Lancet Oncol 21, 1378–1386 (2020).

63. Ho WK, et al. Polygenic risk scores for prediction of breast cancer risk in Asian populations. Genet Med, (2021).

64. Vijg J, Dong X. Pathogenic Mechanisms of Somatic Mutation and Genome Mosaicism in Aging. Cell 182, 12–23 (2020).

