## Supplementary Figures 1-5 and Supplementary Tables 1-8 for "Interplay between mosaic chromosomal alterations and polygenic risk score increases risk of non-small cell lung cancer"

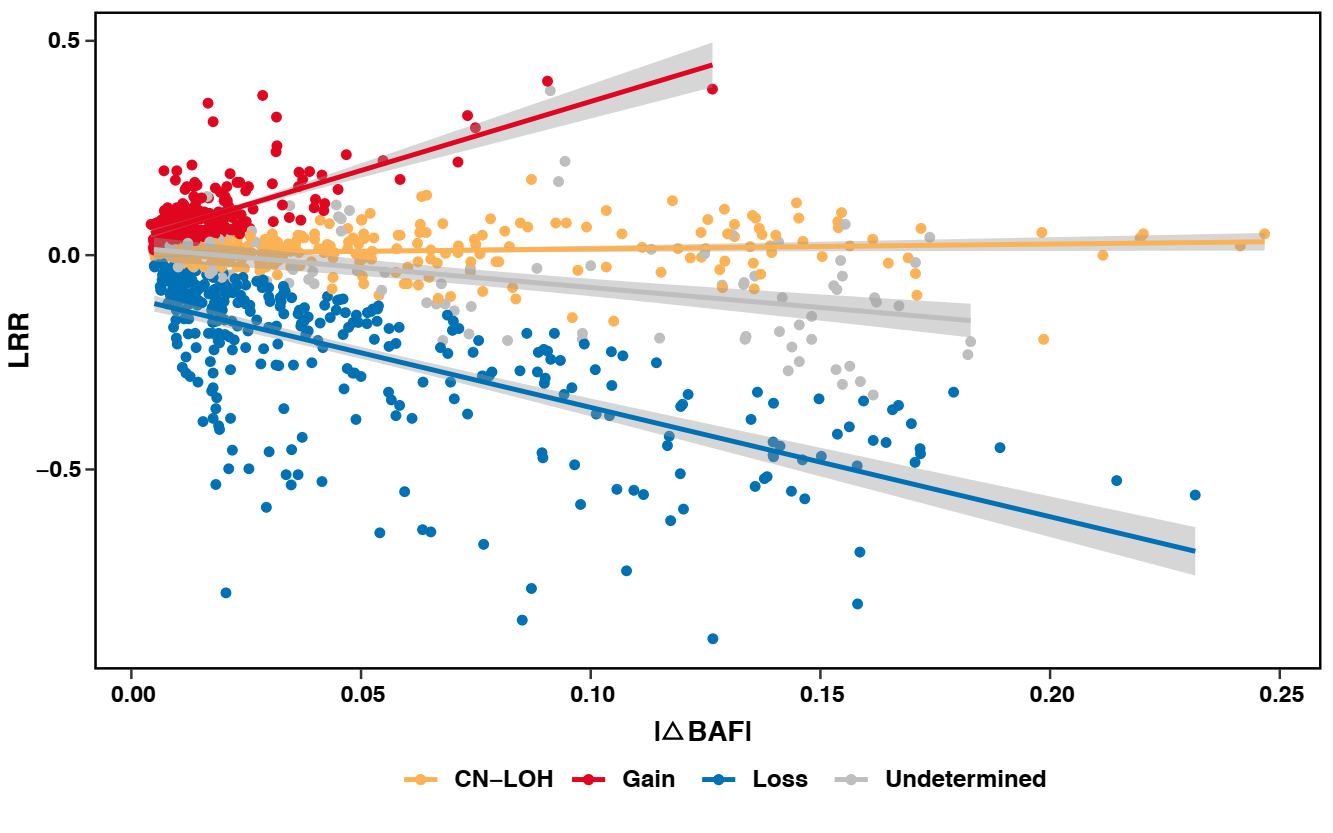
**Supplementary Fig. 1. Classification of mosaic chromosomal alterations (mCAs) as gain, loss or copy-neutral loss of heterozygosity (CN-LOH) events using log-transformed R ratio (LRR) and B allele frequency deviation from 0.5 (|ΔBAF|).** Unclassified events are indicated in grey.

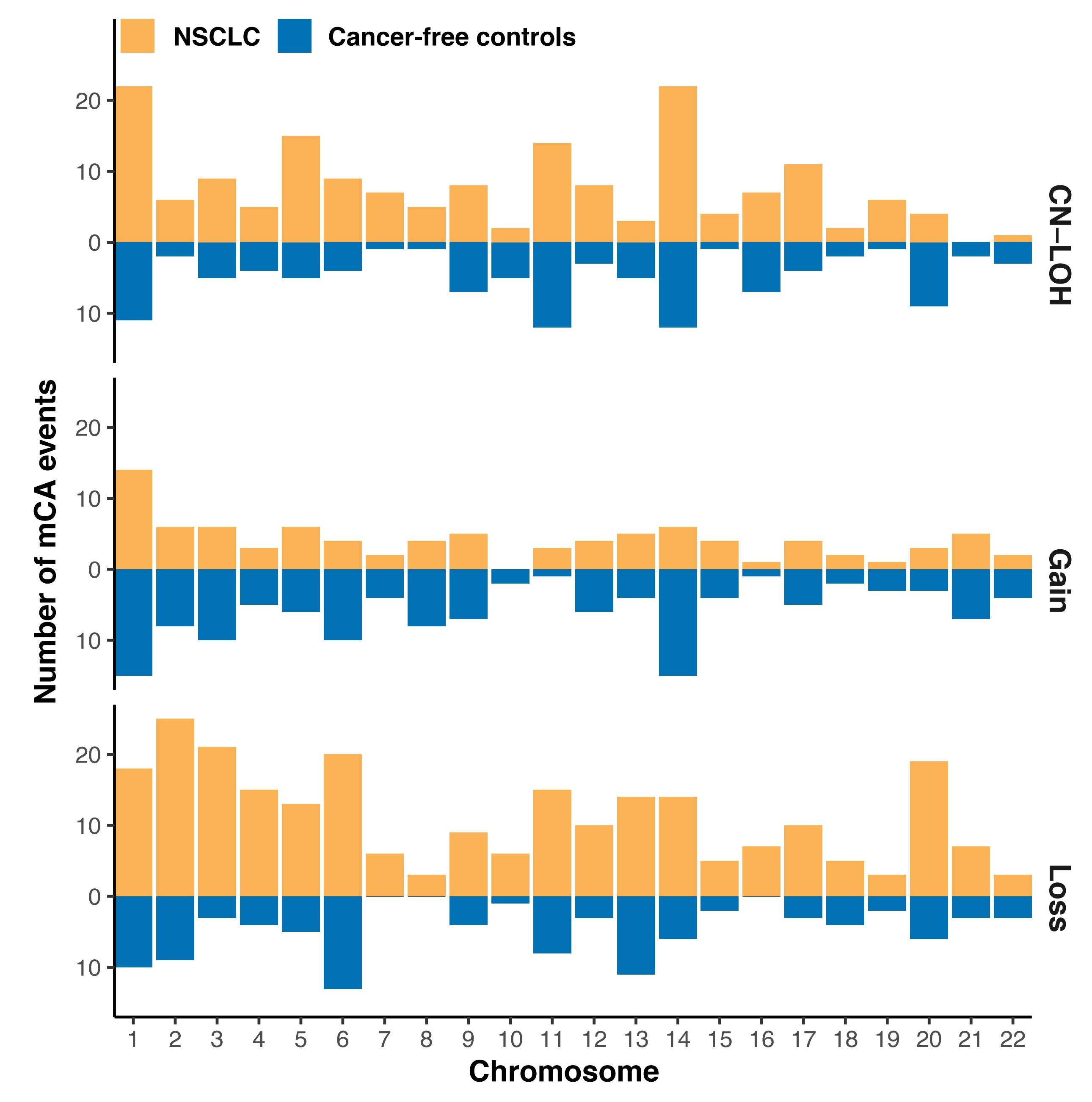

**Supplementary Fig. 2. Distribution of mosaic chromosomal alterations (mCAs) by chromosome.** Abbreviations: CN-LOH, copy-neutral loss of heterozygosity; NSCLC, non-small cell lung cancer.

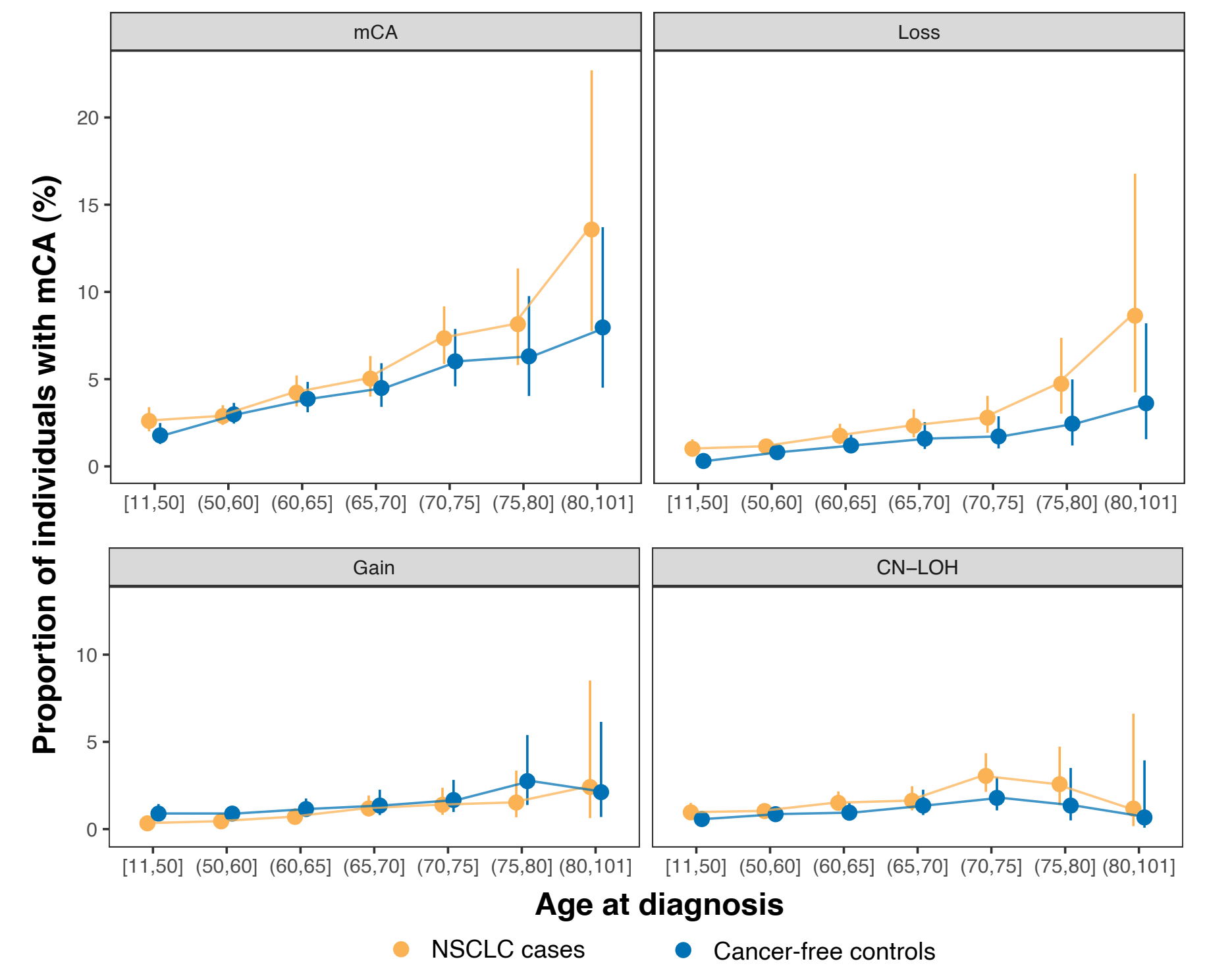
**Supplementary Fig. 3. Association between age and proportion of subjects with detected mosaic chromosomal alterations (mCAs) by copy number alteration types.** Abbreviations: CN-LOH, copy-neutral loss of heterozygosity; NSCLC, non-small cell lung cancer.

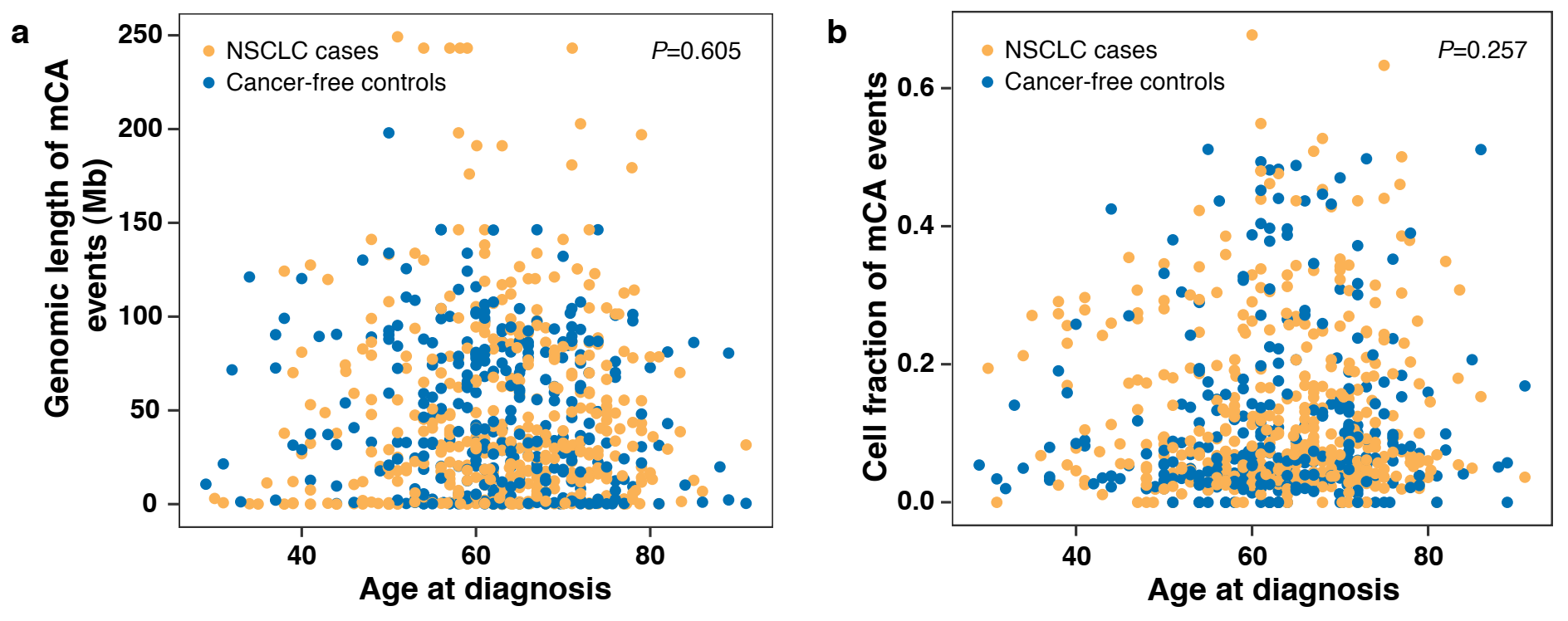

**Supplementary Fig. 4. (a) Association between age and the genomic magnitude of detected mosaic chromosomal alterations (mCAs). (b) Association between age and the cell fraction of detected mosaic chromosomal alterations (mCAs).** Abbreviations: NSCLC, non-small cell lung cancer.

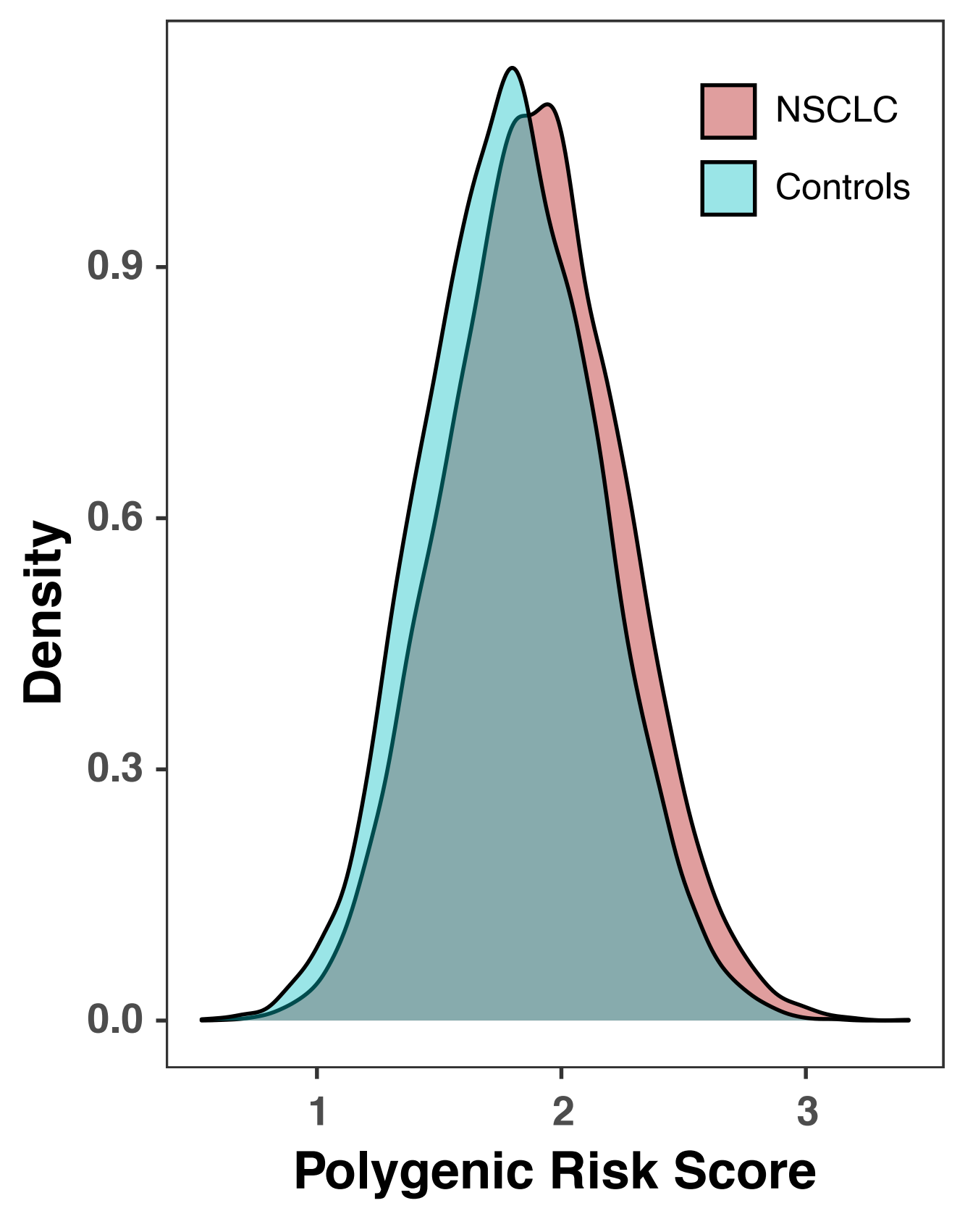
**Supplementary Fig. 5.** **Distribution of polygenic risk score (PRS) in non-small cell lung cancer (NSCLC) patients and cancer-free controls.**

**Supplementary Table 1. Demographic characteristics of the study population.**

|  | **NSCLC cases** | **Cancer-free controls** | ***P*** |
| --- | --- | --- | --- |
|  | **Number (%)** | **Number (%)** |  |
| **Total** | 10248 (100.00) | 9298 (100.00) | - |
| **Age (Mean ± SD)** | 59.01±10.40 | 58.72±10.71 | 0.052 |
| **Sex** |  |  |  |
| Male | 6445 (62.89) | 5871 (63.14) | 0.722 |
| Female | 3803 (37.11) | 3427 (36.86) |  |
| Missing | n/a | n/a |  |
| **Smoking pack-years (PY, Mean ± SD)** | 19.84±27.18 | 11.34±18.90 | **1.01E-141** |
| **Smoking status** |  |  |  |
| Nonsmokers | 5164 (50.39) | 5454 (58.66) | **8.24E-31** |
| Smokers | 5073 (49.50) | 3841 (41.31) |  |
| Missing | 11 (0.11) | 3 (0.03) |  |
| **Histology** |  |  | - |
| Lung adenocarcinoma | 6839 (66.73) | - |  |
| Lung squamous cell carcinoma | 2704 (26.39) | - |  |
| Other^a^ | 705 (6.88) | - |  |
| **Source** |  |  | **1.21E-18** |
| Beijing | 2155 (21.03) | 2035 (21.89) |  |
| Nanjing | 4149 (40.49) | 3198 (34.39) |  |
| Zhongshan | 3944 (38.49) | 4065 (43.72) |  |
| Missing | n/a | n/a |  |

*P* value in bold indicates statistically significant (*P* < 0.05).

a^ The remaining cases include other histological subsets of non-small cell lung cancer, such as large cell carcinoma, mixed histology, etc.

**Supplementary Table 2. Detailed information for the variants used for polygenic risk score (PRS) calculation.**

| **SNP ID** | **CytoBand** | **Position^a^** | **Allele** | | **Ln (OR)^b^** | **# of Risk Alleles** | **# of Risk Alleles*Ln(OR)** |
| --- | --- | --- | --- | --- | --- | --- | --- |
|  |  |  | **Effect Allele** | **Other Allele** |  |  |  |
| rs17038564 | 2p14 | 65496058 | G | A | 0.102 | 0.00 | 0.000 |
| rs2293607 | 3q26.2 | 169482335 | T | C | 0.117 | 1.00 | 0.117 |
| rs11375254 | 3q28 | 189343242 | T | TA | 0.190 | 1.96 | 0.372 |
| rs13167280 | 5p15 | 1280477 | A | G | 0.265 | 1.91 | 0.507 |
| rs401681 | 5p15 | 1322087 | C | T | 0.145 | 2.00 | 0.289 |
| rs2517873 | 6p21.3 | 29875992 | A | G | 0.183 | 0.88 | 0.161 |
| rs3817963 | 6p21.3 | 32368087 | C | T | 0.061 | 1.00 | 0.061 |
| rs1853837 | 6p21.1 | 41497035 | A | C | 0.140 | 0.00 | 0.000 |
| rs5879422 | 6q22.1 | 117784658 | T | TTG | 0.084 | 1.00 | 0.084 |
| rs4236709 | 8p12 | 32410110 | G | A | 0.156 | 1.00 | 0.156 |
| rs10429489 | 9p21.3 | 21787521 | A | G | 0.100 | 0.98 | 0.098 |
| rs35201538 | 9p13.3 | 33422488 | C | CT | 0.095 | 0.00 | 0.000 |
| rs4573350 | 9q33.2 | 124955115 | T | C | 0.090 | 0.00 | 0.000 |
| rs12265047 | 10q25.2 | 114487925 | G | A | 0.134 | 1.98 | 0.265 |
| rs55768116 | 11q23.3 | 118108331 | C | A | 0.118 | 2.00 | 0.237 |
| rs11610143 | 12q13.13 | 52349071 | C | G | 0.065 | 1.94 | 0.126 |
| rs1200399 | 14q13.1 | 35293185 | C | T | 0.100 | 1.00 | 0.100 |
| rs77468143 | 15q21.1 | 49376624 | T | G | 0.077 | 0.00 | 0.000 |
| rs200595745 | 17q24.2 | 65915289 | A | AAATAATAAT | 0.114 | 1.12 | 0.128 |

Abbreviations: OR, odds ratio.

a^ The chromosomal position is shown according to GRCh37.

b^ The estimated weights (lnOR) of the 19 SNPs were derived from our previous study.

**Supplementary Table 3. The prevalence of carriers of autosomal mosaic chromosomal alterations (mCAs) among subgroups.**

| **Variables** | **Group** | **All mCAs** | | |  | **Gain** | | |  | **Loss** | | |  | **CN-LOH** | | |
| --- | --- | --- | --- | --- | --- | --- | --- | --- | --- | --- | --- | --- | --- | --- | --- | --- |
|  |  | **Carriers**  **(N, [%])** | **Non-carriers**  **(N, [%])** | ***P*** |  | **Carriers (N, [%])** | **Non-carriers**  **(N, [%])** | ***P*** |  | **Carriers (N, [%])** | **Non-carriers**  **(N, [%])** | ***P*** |  | **Carriers**  **(N, [%])** | **Non-carriers (N, [%])** | ***P*** |
| **Total** |  | 747 (3.82) | 18799  (96.18) |  |  | 190 (0.97) | 19356  (99.03) |  |  | 277  (1.42) | 19269  (98.58) |  |  | 248  (1.27) | 19298  (98.73) |  |
| Age | <60 yrs | 248 (2.58) | 9375  (97.42) | **1.80E-19** |  | 62  (0.64) | 9561  (99.36) | **3.72E-06** |  | 82  (0.85) | 9541  (99.15) | **3.57E-11** |  | 88  (0.91) | 9535  (99.09) | **1.25E-05** |
|  | ≥60 yrs | 499 (5.03) | 9421  (94.97) |  |  | 128 (1.29) | 9792  (98.71) |  |  | 195  (1.97) | 9725  (98.03) |  |  | 160 (1.61) | 9760  (98.39) |  |
| Sex | Male | 493 (4.00) | 11823  (96.00) | 0.089 |  | 152 (1.23) | 12164  (98.77) | **4.73E-07** |  | 166  (1.35) | 12150  (98.65) | 0.287 |  | 158 (1.28) | 12158  (98.72) | 0.843 |
|  | Female | 254 (3.51) | 6976  (96.49) |  |  | 38  (0.53) | 7192  (99.47) |  |  | 111  (1.54) | 7119  (98.46) |  |  | 90  (1.24) | 7140  (98.76) |  |
| Smoking status | No | 381 (3.59) | 10237  (96.41) | 0.061 |  | 83  (0.78) | 10535  (99.22) | **3.34E-03** |  | 162  (1.53) | 10456  (98.47) | 0.181 |  | 121 (1.14) | 10497  (98.86) | 0.083 |
|  | Yes | 366 (4.11) | 8548  (95.89) |  |  | 107 (1.20) | 8807  (98.80) |  |  | 115  (1.29) | 8799  (98.71) |  |  | 127 (1.42) | 8787  (98.58) |  |

*P* value in bold indicates statistically significant (*P* < 0.05).

Abbreviations: CN-LOH, copy-neutral loss of heterozygosity.

**Supplementary Table 4. Multivariable regression of chronological age, sex, and smoking status on the presence of mosaic chromosomal alterations (mCAs).**

| **Variables** | **Group** | **All mCAs** | | |  | **Gain** | | |  | **Loss** | | |  | **CN-LOH** | | |
| --- | --- | --- | --- | --- | --- | --- | --- | --- | --- | --- | --- | --- | --- | --- | --- | --- |
|  |  | **Carriers (N, [%])** | **OR (95%CI)** | ***P*^a^** |  | **Carriers**  **(N, [%])** | **OR (95%CI)** | ***P*^a^** |  | **Carriers**  **(N, [%])** | **OR (95%CI)** | ***P*^a^** |  | **Carriers**  **(N, [%])** | **OR (95%CI)** | ***P*^a^** |
| Age | <60 yrs | 248 (2.58) | ref | **-** |  | 62  (0.64) | ref | **-** |  | 82  (0.85) | ref | **-** |  | 88 (0.91) | ref | **-** |
|  | ≥60 yrs | 499 (5.03) | 1.99  (1.71, 2.33) | **3.35E-18** |  | 128 (1.29) | 1.92  (1.42, 2.61) | **2.79E-05** |  | 195 (1.97) | 2.36  (1.82, 3.06) | **1.01E-10** |  | 160 (1.61) | 1.78  (1.37, 2.32) | **1.53E-05** |
| Sex | Male | 493 (4.00) | ref | **-** |  | 152 (1.23) | ref | **-** |  | 166 (1.35) | ref |  |  | 158 (1.28) | ref |  |
|  | Female | 254 (3.51) | 0.99  (0.81, 1.20) | 0.892 |  | 38  (0.53) | 0.44  (0.29, 0.67) | **1.25E-04** |  | 111 (1.54) | 1.12  (0.82, 1.52) | 0.488 |  | 90 (1.24) | 1.30  (0.91, 1.86) | 0.142 |
| Smoking status | No | 381 (3.59) | ref | **-** |  | 83  (0.78) | ref | **-** |  | 162 (1.53) | ref |  |  | 121 (1.14) | ref |  |
|  | Yes | 366 (4.11) | 1.12  (0.93, 1.35) | 0.246 |  | 107 (1.20) | 0.99  (0.71, 1.39) | 0.960 |  | 115 (1.29) | 0.88  (0.65, 1.20) | 0.413 |  | 127 (1.42) | 1.46  (1.04, 2.05) | **0.031** |

*P* value in bold indicates statistically significant (*P* < 0.05).

Abbreviations: CN-LOH, copy-neutral loss of heterozygosity.

a^ Status of mCA event (Y/N) ~ Age + Sex + Smoking pack-years.

**Supplementary Table 5. Stratification analysis for associations between autosomal mosaic chromosomal alterations (mCAs) and risk of non-small cell lung cancer (NSCLC).**

| **Variables** | **Group** | **Num** | **All mCAs** | | | **Gain** | | | **Loss** | | | **CN-LOH** | | |
| --- | --- | --- | --- | --- | --- | --- | --- | --- | --- | --- | --- | --- | --- | --- |
|  |  |  | **OR (95% CI)^a^** | ***P*^a^** | ***P*_het_** | **OR (95% CI)^a^** | ***P*^a^** | ***P*_het_** | **OR (95% CI)^a^** | ***P*^a^** | ***P*_het_** | **OR (95% CI)^a^** | ***P*^a^** | ***P*_het_** |
| Age | <60 yrs | 9623 | 1.13 (0.87, 1.48) | 0.352 | 0.771 | 0.52 (0.30, 0.90) | **0.020** | 0.195 | 2.05 (1.27, 3.32) | **3.52E-03** | 0.479 | 1.26 (0.81, 1.96) | 0.311 | 0.726 |
|  | ≥60 yrs | 9920 | 1.19 (0.98, 1.44) | 0.077 |  | 0.81 (0.56, 1.17) | 0.257 |  | 1.67 (1.23, 2.27) | **1.13E-03** |  | 1.39 (0.98, 1.97) | 0.062 |  |
| Sex | Female | 7230 | 1.17 (0.90, 1.52) | 0.231 | 0.953 | 0.34 (0.16, 0.72) | **4.87E-03** | **0.032** | 1.80 (1.21, 2.70) | **4.03E-03** | 0.879 | 1.43 (0.92, 2.23) | 0.110 | 0.762 |
|  | Male | 12316 | 1.18 (0.97, 1.44) | 0.091 |  | 0.84 (0.60, 1.19) | 0.333 |  | 1.73 (1.23, 2.43) | **1.52E-03** |  | 1.31 (0.93, 1.86) | 0.123 |  |
| Smoking status | Yes | 8914 | 1.21 (0.97, 1.51) | 0.097 | 0.529 | 0.80 (0.54, 1.18) | 0.261 | 0.212 | 1.68 (1.11, 2.55) | **0.014** | 0.920 | 1.38 (0.94, 2.04) | 0.098 | 0.875 |
|  | No | 10618 | 1.10 (0.89, 1.35) | 0.400 |  | 0.53 (0.33, 0.87) | **0.011** |  | 1.73 (1.25, 2.40) | **1.07E-03** |  | 1.33 (0.91, 1.93) | 0.139 |  |
| Histology | AD | 16137 | 1.22 (1.03, 1.44) | **0.024** | 0.903 | 0.68 (0.48, 0.98) | **0.037** | 0.548 | 1.81 (1.37, 2.39) | **2.65E-05** | 0.779 | 1.34 (0.99, 1.80) | 0.056 | 0.524 |
|  | SC | 12002 | 1.19 (0.93, 1.53) | 0.168 |  | 0.82 (0.51, 1.33) | 0.428 |  | 1.69 (1.12, 2.54) | **0.012** |  | 1.58 (1.04, 2.40) | **0.032** |  |

*P* value in bold indicates statistically significant (*P* < 0.05).

Abbreviations: CN-LOH, copy-neutral loss of heterozygosity; OR, odds ratio.

a^ NSCLC (case/control) ~ Status of mCA event (Y/N) +Age + Sex + Smoking pack-years + DNA source + 10 Principal components.

**Supplementary Table 6. Associations between autosomal mosaic chromosomal alterations (mCAs) and non-small cell lung cancer risk by genomic location.**

| **Type** | **Ncase^a^** | **Nctrl^a^** | **OR (95% CI)^b^** | ***P*^b^** |
| --- | --- | --- | --- | --- |
| **Telometric** | 258 | 214 | 1.10 (0.92, 1.32) | 0.309 |
| Gain (Telometric) | 58 | 87 | 0.61 (0.44, 0.85) | **3.48E-03** |
| Loss (Telometric) | 91 | 37 | 2.24 (1.53, 3.29) | **3.57E-05** |
| CN-LOH (Telometric) | 98 | 84 | 1.06 (0.79, 1.43) | 0.676 |
| Undetermined (Telometric) | 26 | 13 | 1.82 (0.94, 3.55) | 0.077 |
| **Interstitial** | 184 | 118 | 1.42 (1.13, 1.80) | **3.01E-03** |
| Gain (Interstitial) | 15 | 20 | 0.68 (0.35, 1.34) | 0.267 |
| Loss (Interstitial) | 100 | 60 | 1.52 (1.10, 2.10) | **0.011** |
| CN-LOH (Interstitial) | 61 | 14 | 3.98 (2.22, 7.11) | **3.27E-06** |
| Undetermined (Interstitial) | 28 | 33 | 0.77 (0.47, 1.28) | 0.320 |
| **Whole chromosome events** | 21 | 10 | 1.92 (0.90, 4.07) | 0.091 |
| Gain (Whole chromosome events) | 12 | 9 | 1.22 (0.51, 2.89) | 0.657 |
| Loss (Whole chromosome events) | 7 | 1 | 6.39 (0.79, 51.71) | 0.082 |
| CN-LOH (Whole chromosome events) | 3 | 0 | 96219.64 (0.00, >10^100^) | 0.920 |
| Undetermined (Whole chromosome events) | 2 | 1 | 1.82 (0.17, 20.11) | 0.623 |
| **Centromeric** | 7 | 3 | 2.13 (0.55, 8.23) | 0.273 |
| Gain (Centromeric) | 1 | 2 | 0.46 (0.04, 5.03) | 0.522 |
| Loss (Centromeric) | 6 | 1 | 5.47 (0.66, 45.45) | 0.115 |

*P* value in bold indicates statistically significant (*P* < 0.05).

Abbreviations: CN-LOH, copy-neutral loss of heterozygosity; OR, odds ratio.

a^ Number of mCA carriers among non-small cell lung cancer cases and cancer-free controls respectively.

b^ NSCLC (case/control) ~ Status of mCA event (Y/N) + Age + Sex + Smoking pack-years + DNA source + 10 Principal components.

**Supplementary Table 7. Associations between cell fractions of autosomal mosaic chromosomal alterations (mCAs) and risk of non-small cell lung cancer (NSCLC).**

|  | **NSCLC cases** | **Controls** | **Univariable Model** | | **Multivariable Model^a^** | |
| --- | --- | --- | --- | --- | --- | --- |
|  | **Carriers, N (%)** | **Carriers, N (%)** | **OR (95% CI)** | ***P*** | **OR (95% CI)** | ***P*** |
| Total mCAs | 418 (4.08%) | 329 (3.54%) | 3.92 (1.62, 9.53) | **2.54E-03** | 4.28 (1.70, 10.79) | **2.04E-03** |
| CN-LOH | 153 (1.49%) | 95 (1.02%) | 16.07 (2.82, 91.61) | **1.77E-03** | 11.27 (1.83, 69.42) | **9.04E-03** |
| Gain | 79 (0.77%) | 111 (1.19%) | 0.19 (0.01, 3.06) | 0.245 | 0.37 (0.02, 7.09) | 0.511 |
| Loss | 179 (1.75%) | 98 (1.05%) | 4.14 (1.34, 12.84) | **0.014** | 4.81 (1.49, 15.51) | **8.55E-03** |

*P* value in bold indicates statistically significant (*P* < 0.05).

Abbreviations: CN-LOH, copy-neutral loss of heterozygosity; OR, odds ratio.

a^ NSCLC (case/control) ~ Cell fraction of mCA event +Age + Sex + Smoking pack-years + DNA source + 10 Principal components.

| **PRS^d^** | **Multiplicative interaction^a^** | | **Additive interaction^b^** | | | | | |
| --- | --- | --- | --- | --- | --- | --- | --- | --- |
|  |  |  | **All mosaic loss events** | | **mosaic loss with cell fraction <5%** | | **mosaic loss with cell fraction≥5%** | |
|  | **OR (95%CI)** | ***P* value** | **RERI^c^ (95%CI)** | **AP^c^ (95%CI)** | **RERI^c^ (95%CI)** | **AP^c^ (95%CI)** | **RERI^c^ (95%CI)** | **AP^c^ (95%CI)** |
| Intermediate genetic risk | 2.33 (1.08, 5.00) | **0.030** | 0.65 (-0.49, 1.85) | 0.28 (-0.31, 0.53) | -0.03 (-2.55, 2.6) | -0.02 (-2.77, 0.41) | 1.21 (-2.01, 6.65) | 0.33 (-1.24, 0.57) |
| High genetic risk |  |  | 3.72 (1.13, 7.87) | 0.61 (0.25, 0.74) | 0.85 (-0.52, 2.28) | 0.34 (-0.31, 0.59) | 5.26 (1.54, 12.10) | 0.69 (0.31, 0.80) |

**Supplementary Table 8. Analyses for interaction between polygenic risk score (PRS) and autosomal mosaic loss abnormalities.**

*P* value in bold indicates statistically significant (*P* < 0.05).

Abbreviations: AP, attributable proportion; RERI, relative excess risk due to interaction; OR, odds ratio.

a^ NSCLC (case/control) ~ PRS (continuous variable) + mosaic loss (Y/N) + Age + Sex + Smoking pack-years + DNA source + 10 Principal components + PRS × mCA (Y/N)

b^ NSCLC (case/control) ~ PRS (categorical variable) + mosaic loss (Y/N) + Age + Sex + Smoking pack-years + DNA source + 10 Principal components + PRS (categorical variable) × mCA (Y/N)

c^ The non-carriers of mosaic loss events within the lowest genetic risk (the first quartile) group was set as the reference categories.

d^ Genetic risk was categorized into low (the first quartile), intermediate (25%~75%) and high (the upper quartile) according to distributions of PRS among cancer-free controls.
